# Feasibility of Continuous Distal Body Temperature for Passive, Early Pregnancy Detection

**DOI:** 10.1101/2021.08.19.21262306

**Authors:** Azure Grant, Benjamin Smarr

## Abstract

The majority of American women become aware of pregnancy ~3-7 weeks after conception, and all must seek testing to confirm their pregnant status. The delay between conception and awareness is often a time in which contraindicated behaviors take place. However, there is long standing evidence that passive, early pregnancy detection may be possible using body temperature. To address this possibility, we analyzed 30 individuals’ continuous distal body temperature (DBT) in the 180 days surrounding self-reported conception in comparison to self-reported pregnancy confirmation. Features of DBT nightly maxima changed rapidly following self-reported conception, reaching uniquely elevated values after a median of 5.5 ± 3.5 days, whereas individuals reported a positive pregnancy test result at a median of 14.5 ± 4.2 days. Together, we were able to generate a retrospective, hypothetical alert a median of 9 ± 3.9 days prior to the date at which individuals received a positive pregnancy test. Continuous temperature-derived features can provide early, passive indication of pregnancy onset. We propose these features for testing and refinement in clinical settings, and for exploration in large, diverse cohorts. The development of pregnancy detection using DBT may reduce the delay from conception to awareness and increase the agency of pregnant individuals.

## Introduction

### Why is early pregnancy detection needed?

Automated, early pregnancy detection might substantially improve maternal and fetal health. Although population reports of pregnancy testing behavior are sparse^1–3^, available cohorts indicate that most American women confirm pregnancy about 3 to 5 weeks after conception, and that nearly a quarter do so later than 5 weeks^1^. The cost of delayed pregnancy awareness is great: adoption of appropriate pregnancy habits is delayed^4,5^ in the earliest, most fragile developmental window^6–9^. Moreover, unplanned pregnancy occurs at rates of ~50%^10–13^, and is associated with higher likelihood of adverse exposures^4,5,14,15^, increased maternal morbidity and mortality^16,17^, preterm birth and low childhood weight^18–21^, elevated risk of birth defects^22,23^, and poorer maternal psychological health^24,25^. As the effective window for emergency contraception is ~120 h at most^26–29^, and as safe abortion access continues to be limited around the world^16,30^, delayed confirmation poses considerable risks to pregnant individuals^16,17^. Early, passive pregnancy detection could increase the agency of a pregnant individual, speed adoption of pregnancy-safe behaviors (e.g., avoidance of environmental risk factors^7^, cessation of alcohol consumption^8,31^ or drug use^32^), or provide the choice to discontinue a pregnancy at an earlier gestational age^33^.

### What is the current state of pregnancy detection?

The development of automated tools has been limited by our low temporal resolution understanding of somatic changes in early pregnancy, and an historical reliance on single time-point hormone samples. Current clinical or over-the-counter (OTC) pregnancy tests rely on serum or urine measurements of human chorionic gonadotropin (hCG), and claim to detect elevated hCG with 99% accuracy at approximately the date of missed menses^34^; however, independent studies place most at-home test kit accuracies far lower, even weeks after missed menses^34–37^. Additionally, as pregnancy test efficacy is measured in days relative to expected missed period (i.e., about 12-17 days after conception^38^), the number of days post-conception that a test becomes accurate is both difficult to estimate^38^ and likely more variable for the ~1/3 of women with irregular or long menstrual cycles^39^. Although OTC testing provides a potentially early indicator, most individuals who seek testing do not do so until weeks after a positive test may be obtained^1–3^. For unplanned pregnancies, this delay may be even more prolonged.

### Why do we suspect that temperature can advance pregnancy testing?

There is reason to believe that passively measurable outputs with direct mechanistic ties to female reproductive physiology, such as continuous body temperature, could advance pregnancy testing^40,41^. A growing number of wearable sensors offer non-invasive, continuous body temperature measures^40,42–44^. Such timeseries, if appropriately measured and analyzed^42,45,46^, provide a window onto reproductive events with great temporal granularity, and create an opportunity for precise mapping of patterns of change in pregnancy^40^.

Elevation of basal body temperature (BBT) during pregnancy, and its putative relationship to progesterone, has been recognized for about a century^47–49,50–68^. However, BBT has proven too imprecise and difficult to collect for development as a reliable method of either luteinizing hormone surge^69–71^, ovulation^70–74^, or early pregnancy detection^75–78^. Despite this, community groups^79^ frequently adapt BBT-based methods of ovulatory cycle tracking (i.e., the sympto-thermal method) in an attempt to detect pregnancy onset^80,81^. Although efforts have been made to automate the collection of BBT or similar metrics, they have focused largely on proception or contraception rather than on pregnancy detection^82–85^. However, methods of continuous (as opposed to once per day) collection of temperature are a promising alternative that appears to provide large gains in information about reproductive status by examining not only levels but carefully selected patterns of temperature^40–42,86,87^.

For example, murine elevation of continuous body temperature can be used to detect pregnancy onset within ~12 h of conception^41^. Subsequent investigations in humans revealed continuous temperature-based predictors of the preovulatory LH surge^42^, as well as prediction and detection of sickness^88^. Based on this growing body of work and on community reports, we hypothesized that features extracted from continuous distal body temperature at the finger (DBT) could provide early indicators of pregnancy. Here we report results from retrospective wearable temperature data from 3 months prior to 3 months after self-reported conception and compared this to self-reported dates of OTC or clinically confirmed pregnancy.

## Results

### Descriptive Statistics

#### Distal Body Temperature Could Cut the Time to Pregnancy Awareness by Over Two Thirds in This Feasibility Cohort

Features of DBT exhibited unique elevations following self-reported date of conception (**See: Methods**) in all 30 cases, as well as reproducing previous reports of apparent ovulatory cycles^42,43^ (**Fig 1**). Unique DBT elevations were used to create a Retrospective Hypothetical Alert (RHA) (**See: Methods**) that was triggered before standard positive pregnancy test results in 29/30 cases (**Fig 1, Table 1**). This is illustrated in Figure 1 as the transition from unconfirmed pregnancy following conception (red) into RHA-tagged pregnancy (blue), which precedes the transition back to grey that occurs upon confirmatory testing. RHA occurred a median duration of 5.5 ± 3.5 days after reported conception, whereas reported confirmation using a standard pregnancy test occurred after a median of 14.5 ± 4.2 days. The RHA-confirmed days occurred significantly earlier than pregnancy test confirmation (p=1.05*10^− 8^). To provide context for the potential utility of such passively-generated alerts, we highlighted comparison to average pregnancy confirmation dates, including the U.S. population at large, as well as two populations that are at higher risk of pregnancy complications, “Black and Hispanic”^89,90^ and “Teen”^1,16^. Additionally, we highlighted the estimated 23% of U.S. pregnancies which are not confirmed by approximately 5 weeks after conception^1^.

**Table 1.** Age distribution and confirmatory testing types for this cohort.

|  |  |  |  |  |
| --- | --- | --- | --- | --- |
| Age Range (%) | 25-29 (10%) | 30-34 (50%) | 35-39 (23%) | 40-44 (17%) |
| Method of Pregnancy Confirmation (%) | OTC Test (80%) | In-Clinic Test (7%) | IVF Initiated Pregnancy (13%) |  |

**Figure 1.**
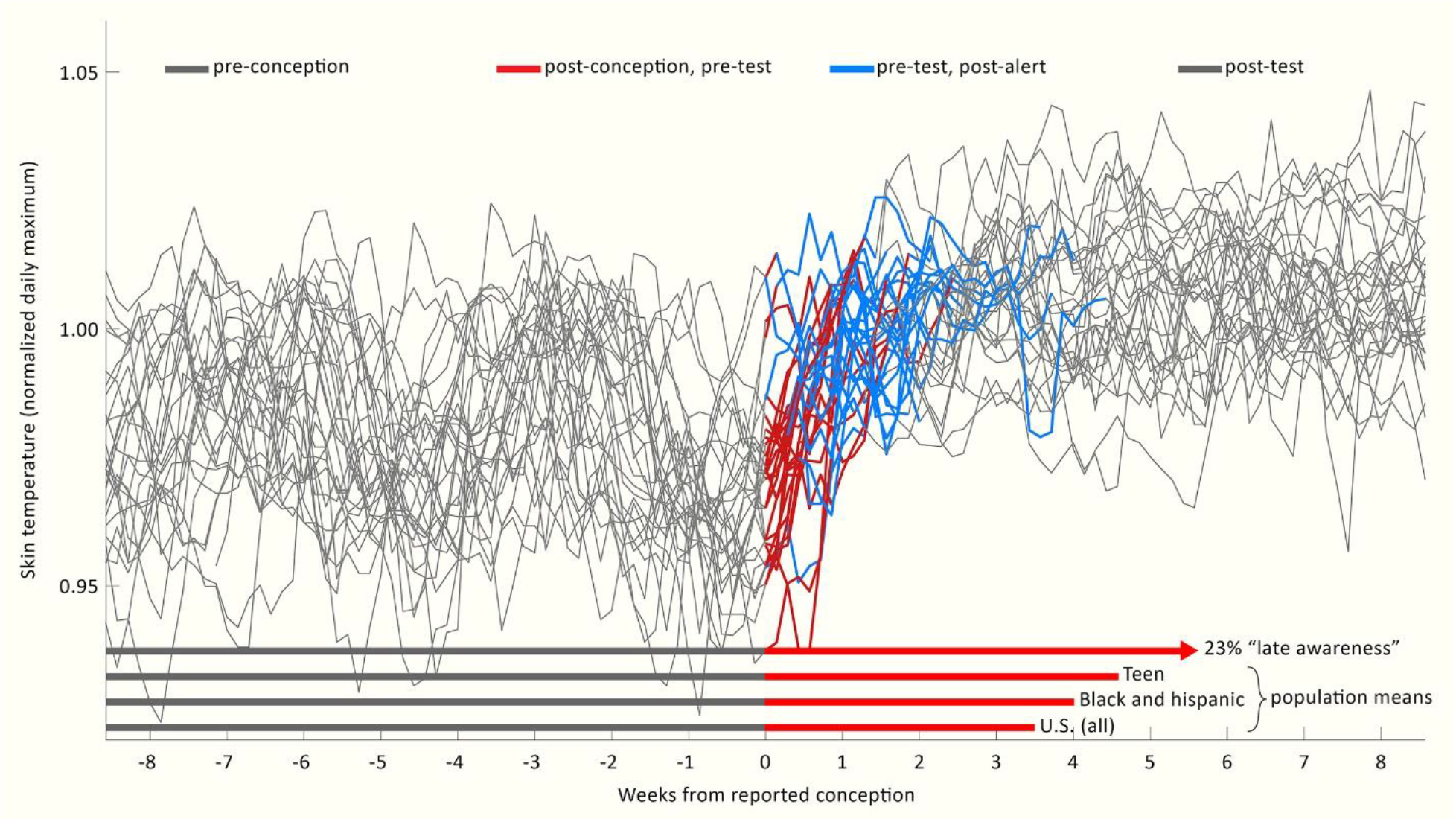
Nightly temperatures allow generation of retrospective hypothetical alerts in advance of pregnancy confirmation by standard tests. Individuals’ nightly maximum temperatures, aligned to the date of reported conception (grey) for all 30 participants. Red: Time between reported conception and reported test confirmation is shown in red unless (Blue) an RHA occurred for that individual, at which point red is replaced by blue. At the date of test confirmation, records return to grey. Bottom: red vectors represent mean time from conception to confirmation for: the U.S. population (3.5 weeks); vulnerable sub populations of “Black and Hispanic” (3.9 weeks) and “Teen” (4.6 weeks), and the 23% of the U.S. population reporting “late confirmation” of more than 5 weeks after conception. Epidemiological data visualized from^1^.

#### Nightly Maxima Uniquely Reflect Early Pregnancy

Features of DBT were extracted in order to detect pregnancy onset. Raw DBT (**Fig 2A**) was bimodally distributed (**Fig 2B**). DBT oscillated between relatively labile, lower daytime or waking values, and relatively higher, more tightly distributed nighttime or sleeping values (**Fig 2B,D**)^42,88,91^. Comparison of the daily highs (**Fig 2C, top**) to the daily lows (**Fig 2C, bottom**) confirmed that putative ovulatory cycles and pregnancy onset were more apparent as changes in nightly levels. Representative nighttime temperature (**See: Methods** for feature derivation) from each participant for each night confirmed the ability to detect roughly-monthly cycles preceding reported conception, as well as a stereotyped rise following conception (**Fig 2D,E**). Nightly maxima exhibited values that were statistically elevated over previous luteal phase peaks by 6 days after reported conception (χ^2^=108, p=3.16*10^−18^, L vs. C +6 days p=0.018, L +7 days and onward p< 1*10^−4^) (**Fig 3A, 3C** light blue panel).

**Figure 2.**
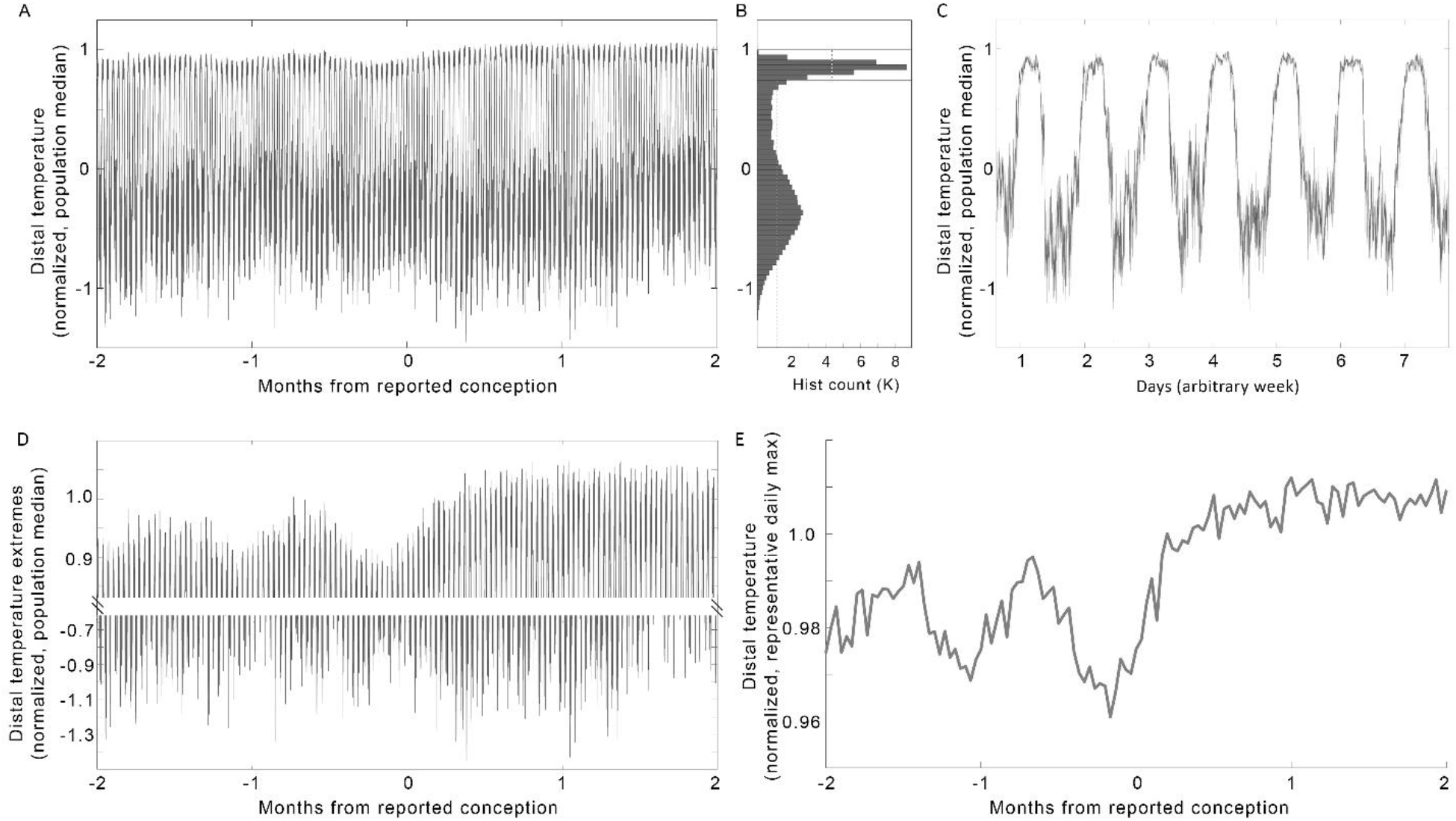
Raw temperature must be transformed to clearly reveal pregnancy onset trends. (A) Median raw minutely temperature records from all 30 individuals, from 2 months before to 2 months after reported conception. (B) Histogram of pre-conception temperatures in (A) revealed a bimodal distribution, reflecting day-night oscillations, as seen more clearly in a zoom of one arbitrary week (C). (D) Data from A, zoomed to highlight the nightly highs (top) and daily lows (bottom). (E) Population median of the nightly temperature maxima (see: **Methods**) enhanced the clarity of the patterns reflecting putative ovulatory cycles and pregnancy onset contained within DBT.

**Figure 3.**
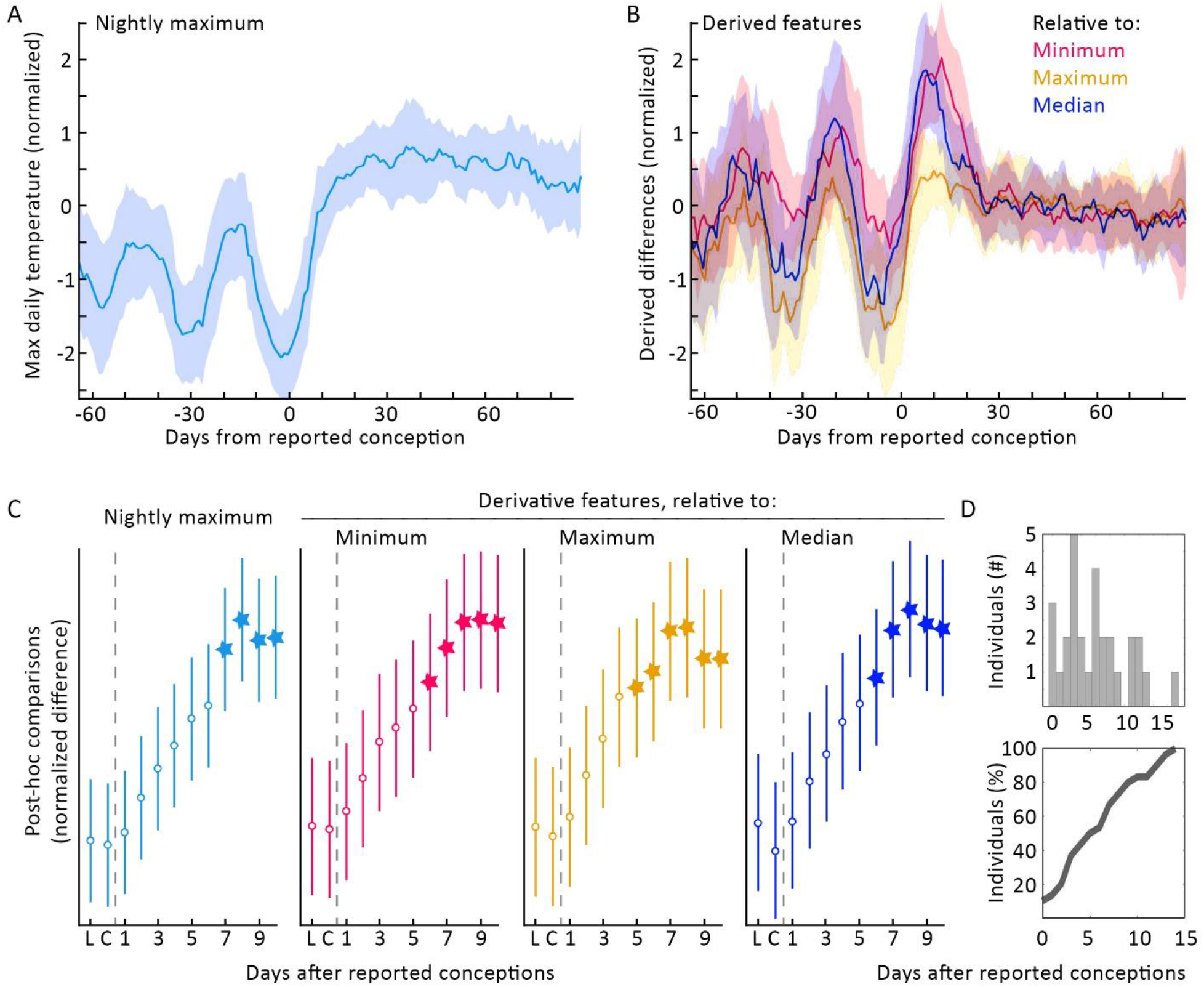
Transformations of nightly temperature maxima enhanced the change seen at pregnancy onset and allowed more efficient RHAs for all 30 participants. (A) Median nightly temperature maxima ± MAD (see: **Methods**). (B) Overlay of the three features derived from comparison to individuals’ historical medians (blue), minima (magenta), and maxima (gold) (see: **Methods**). Post-hoc tests of the difference between putative luteal-phase peaks, and the difference between putative-luteal phase peaks (“L”) and days from conception (“C”), indicated significant elevation of all features from post-conception days 5-6 onward. The derived feature “Maximum” reached statistical significance faster (5 days) than did the other features (6 days). Linear (D, top) and cumulative (D, bottom) histograms of days on which these features generated an RHA (see **Methods**).

#### Normalizing Nightly Maxima to Within-Individual History Improved Speed of Pregnancy Detection

In order to maximize contrast at transitions from follicular to luteal phases, and from follicular phases to pregnancies, we generated second order features normalized to the average historical length of the follicular phase (17 days^39^) (**See Methods** for discussion of normalization window). Normalization of an individual’s nightly maxima to the previous 17 days’ history of minima (**Fig 3B, 3C** red), maxima (**Fig 3B, 3C** gold), and medians (**Fig 3B, 3C** dark blue) exhibited variably faster within-individual statistical elevations above luteal highs. The derived feature comparing each nightly maximum to historical nightly maxima achieved significance 5 days after reported conception (χ^2^=72.2, p=4.62*10^−11^, L vs. C +6 days p=0.014, L +7 days and onward p< 3*10^−4^), whereas those comparing to historical minima and medians reached statistically significant elevation 6 days after reported conception (relative to historical minima: χ^2^=78.00, p=3.59*10^−12^, L vs. C +6 days p=0.034, L +7 days and onward p< 1.60*10^−3^; relative to historical medians: χ^2^=91.0, p=1.08*10^−14^, L vs. C +6 days p=0.040, L +7 days and onward p< 4*10^−3^). Although multiple features reached group significance, on the same day, different individuals reached each feature at different times, as shown in the histogram of RHAs (**Fig 3D**).

#### Case Study: Similarity Among Mouse and Human Temperatures Across Pregnancy

We previously published the use of features derived from continuous core body temperature to detect pregnancies in mice, and to monitor pregnancies through delivery^41^. Here we highlighted a comparison of a mouse and a human mother across gestation (**Fig 4**) in order to support future investigations of continuous body temperature patterns beyond conception. Despite obvious differences in gestational length, body size, and the species’ reproductive physiology, we noted several striking visual similarities between the two pregnancies. Both exhibited ovulatory cycles, followed by a steep rise in temperature at conception, and a long slow decrease in temperature until around the 3^rd^ trimester. Then, large “hills” appeared in the temperature trajectory, followed by another steep rise preceding delivery. The consistency and mechanisms of these changes are beyond the scope of this manuscript. We hope that sharing this visual inspection is sufficient support the apparent existence of these conserved patterns in DBT and support more research in the field.

**Figure 4.**
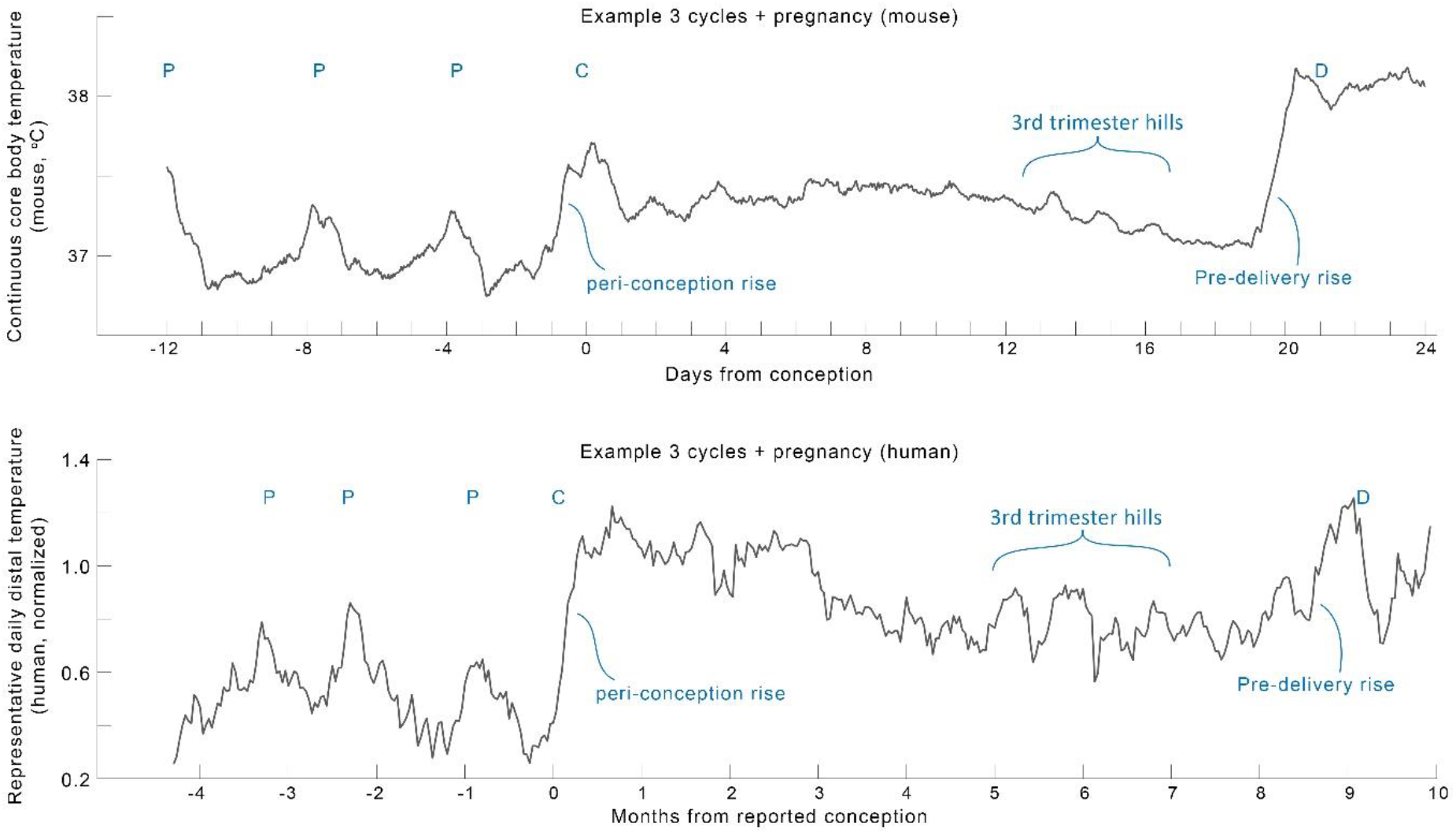
Comparison of one example mouse (top, from^41^) and human (bottom) pregnancy profile revealed many apparently parallel structures in continuous temperature in need of deeper mechanistic investigation. To highlight trends over daily variance, both profiles show moving means with windows of mouse (24 hours on minutely data) and human (7 days on nightly temperature maxima) data. “P”: peaks of putative ovulatory cycles, “C”: conception, “D”: delivery.

## Discussion

The present study supports the feasibility of passive, early pregnancy detection using continuous DBT. In this cohort, nightly temperature maxima rose rapidly in early pregnancy and reached uniquely high values an average of 5.5 days after self-reported conception. These features enabled generation of RHAs for all individuals, which illustrated the potential to cut time to pregnancy awareness by 2/3 as compared to reported use of pregnancy tests^1^. Additional analysis of large, diverse cohorts is necessary to understand population variability in DBT patterns and to appropriately refine the features described here before attempting to create prospective alerts. Furthermore, preliminary comparison to published murine data suggests that additional features of DBT may be useful for pregnancy applications beyond conception.

The potential utility of temperature measurement for pregnancy detection was published nearly 100 years ago^47,48,92^. Van de Velde reported a sharp rise in daily oral basal body temperature (BBT) in early pregnancy^47,48^, and Buxton and Atkinson subsequently postulated its dependence on elevation of progesterone^49^. Amazingly, even finger temperature was proposed in 1949 as a method of pregnancy monitoring^93^ (although the authors were not able to locate primary follow-up studies^94–98^). Despite this, oral thermometry has failed to develop as a trusted tool for early detection^75–77,99,78^. Why might continuous distal temperature improve upon historical uses of oral temperature? By definition, continuous measurement allows more information to be captured per unit time. Practically, continuous DBT provides a much wider range of potential features associated with adaptation to a pregnancy than does BBT^40,100^, such as extracting the nightly maxima studied here. At the periphery of the body, temperature changes greatly by sleep-wake status^91^, circadian and ultradian phase^101,102^. Moreover, DBT is temporally coupled to physiological systems that rise in output across the first trimester of pregnancy, such as progesterone^103,104^, night time heart rate^105^, insulin activity^106–108^, luteinizing hormone^42^, and cortisol^109,110^.

This additional temporal structure, and more thoroughly mapped relation to hormonal output, allows intentional selection of timeseries features that reflect physiological changes. In this case, sleeping temperature measurement provides a relatively unperturbed state (akin to the state that first morning BBT roughly attempts to capture), in which an individual’s behaviors (e.g., exercise, outdoor environment) may have a more limited effect on temperature level and pattern. It is possible that this relative absence of behavioral contributions enables endocrine contributions to the DBT signal to take precedence^111,112^. As pregnancy is associated with substantial changes to hormone levels and thermoregulation, as well as cardiovascular remodeling, future research is needed to confirm that relationships between temperature and hormonal status hold in pregnant individuals.

Notably, the authors were unable to find high temporal resolution descriptions of peripheral physiology around the time of conception. It is possible that the present lack of tools for conception detection makes the study of very early natural pregnancies challenging. If features such as those described here persist in larger cohorts, then continuous DBT may enable more precise study of the physiological changes associated with conception, including continuous glucose patterning^106,107^, salivary and urinary hormone output^113,114^, early cardiovascular remodeling^115^. Additionally, continuous data may allow definition of tighter critical windows for the impacts from maternal and environmental influences^8^. Much remains to be learned about physiological diversity in continuous DBT, and there are likely myriad strategies for generating predictive features. The present study supports the hypothesis that wearable devices can be used to map this physiological diversity and identify features that provide useful information on an individualized basis.

Such tools are not without risk. False positives could result in unnecessary psychological stress or unwarranted consumption of emergency contraception^116^, whereas false negatives could lead to a lack of caution in a true early gestation. Additionally, this study did not differentiate pregnancies that go on to have second trimester miscarriage, stillbirth, or other complications, and future work is needed to uncover if unique conception profiles are associated with these outcomes^47,78^. Moreover, health algorithms have a history of bias and failure to generalize^117^, and our cohort likely represents a relatively affluent and health literate sector of society, which cannot be taken as broadly representative.

Furthermore, retrospective questionnaires like those used in the present study must rely on fallible self-report^118,119^. For instance, self-report of day of conception is prone to error, as many individuals have sex multiple times during the fertile window preceding pregnancy onset^74^. For these reasons, our work supports the need for large, clinical studies on diverse populations to determine which parameters may provide precise indication of conceptive sex, fertilization, and subsequent events. More broadly, our work supports exploration of physiological diversity from timeseries data.

Our analyses of continuous DBT demonstrate the feasibility of passive, early pregnancy screening. Confirmation and augmentation of these features in clinical settings and across large, heterogeneous populations may enable development of wearable tools for pregnancy monitoring. Such tools may increase individuals’ agency and guide pregnancy testing at an earlier gestational age^8^.

## Methods

### Study Design and Data Collection

#### Ethical approval

This study and all experimental protocols were IRB approved by the Office for the Protection of Human Subjects at the University of California, San Diego. All participants gave informed consent. All research was performed in accordance with relevant guidelines and regulations.

#### Participants

All data were collected retrospectively. A study invite was delivered to users of the Oura Ring app via an in-app card which linked users to information about the study, informed consent, and which allowed users to opt-in to share data with the researchers. There were no age or parity restrictions, consistent with the principles of participatory research.

#### Data Collection and Management

Retrospective data had been collected using the Oura Ring (Oura Inc., San Francisco, CA; Oura Health Oy, Ltd., Oulu, Finland). The Oura Ring is a small, wireless sensor worn on the finger. The ring contains 3 thermistors for detection of DBT. DBT is measured 24 h a day at a resolution of once per minute. Raw DBT data are synched from the ring to Oura Ring’s cloud architecture when users open the Oura Ring app on their paired smartphone. This architecture meets CCPA and GDPR privacy and security standards, and provided encrypted, password protected data access to researchers, with the participants’ revocable consent.

In addition to data collected by the Oura Ring, participants provided questionnaire responses. Relevant fields to this manuscript included approximate age at conception (5 year bins), the date the individual believes they conceived, and the date the individual recalls confirming the pregnancy by an over-the-counter or in-lab pregnancy test. Participants could opt out of the study and remove their data from the study pool at any time for any reason. Following data collection, data were anonymized by the researchers for analysis.

#### Inclusion and Exclusion Criteria for Submitted Pregnancies

The participants for this initial study were the first thirty respondents who met the following data quality criteria and questionnaire responses: 1) No data holes > 1 h within the sleeping window in the 60 days surrounding self-reported conception; 2) Positive response for self-reported date of conception; 3) Positive response for self-reported date of pregnancy confirmation via an at-home or in-lab test; 4) Dates provided did not contain obvious typos (e.g., an individual specifying conception on a date that is currently in the future would not be included); 5) Participants did not report a miscarriage following the conception. Note that many pregnancies were ongoing at the time of this study (i.e., individuals were in their second or third trimesters at the time of data collection). This means that we could not rule out the possibility of late miscarriages, still births, or other pregnancy complications within this data set.

### Data Analysis

#### Data Availability and Cleaning

All code and data used in this paper are available at A.G.’s Github^120^. Data were formatted in Excel 2020 and analyses were performed in MATLAB 2021a. Briefly, data were imported from the Oura database into Excel and DBT data were extracted, beginning at midnight 90 days prior to conception and ending approximately 90 days after conception. Data were padded with zeros to obtain a common set of 180 day-long temperature files. Data were cleaned in MATLAB, with any points showing near instantaneous change, as defined by local abs(derivative) > 10^5^ as an arbitrary cutoff set to the median value of the following hour. Very low remaining values, less than 10° C were considered periods of non-wear and were replaced with blanks. Erroneously high values were not observed, with local highs within the physiological range attributable to common activities such as exercise.

#### Temperature Normalization to Pre-Pregnancy Range

Cleaned data were then normalized using representative daily ranges of pre-pregnancy data in order to remove variance associated with individuals with differing average temperatures. Briefly, a representative nightly median was generated for each day from 12am-4am, and a representative daily median was generated for each day from 12pm-4pm. The median of nightly and daily pre-pregnancy values formed the within-individual range to which each participant was normalized. The cooler (daytime) representative value of an individual was subtracted from each data point, and the resultant number divided by the individual’s pre-pregnancy range (representative night value minus representative day value). The resultant normalized temperature was designed such that individuals’ temperature would peak at values of approximately 1.

#### Nightly Temperature Max Evaluation

As finger temperature reaches its maximum values during sleep, and is relatively suppressed during the daytime hours, the median of the hottest 60 minutes of data from a broad window, 8:30pm through 12:00pm the next day, were taken as a representation of maximum temperature during the day. Data were threshold cleaned of sparse high and low outliers, and this nightly maximum variable was saved for each individual for further processing.

#### Nightly Temperature Relative to Features of Estimated Previous Follicular Phase

In order to assess each night’s maximum relative to values in an approximated previous follicular phase (an average of 17 days in large populations^39^), we calculated a rolling past baseline using movmin, movmedian, and movmax functions. We selected 17 days rather than the historical choice of 14 day average follicular phase length in order to impose a stricter threshold for post-conception temperature rise, and in order to encompass the latest data on estimated cycle structure. As we do not have information on individuals’ historical dates of ovulation, we were unable to create personalized follicular phase length normalizations; such personalization will require investigation in future cohorts with measurement of ovulation. We subtracted each of the above nightly maximum values to create a series of personal-history-normalized temperature values, depicted in **Figure 3**. These values can be interpreted as an individual’s hottest sleeping temperatures relative to that individual’s hottest, coldest, or median sleeping values over the prior 17 days. This window was selected based on population reports of typical phase lengths and was grossly intended to improve detection of the contrast between ovulatory cycle phases and conceptions. These final features, nightly max, and nightly max relative to estimated follicular median, maximum, and minimum were normalized by subtracting each individual’s median and dividing by that individual’s median absolute deviation (M.A.D.) before averaging in order to emphasize the pattern of relative change within an individual rather than the absolute y-values of the variables as the key features.

#### Detecting an Individual’s Pregnancy Onset

For each of the above variables - nightly maximum, min-normalized, max-normalized, and median-normalized - the day was specified at which the value of the features exceeded the value of the previous 2 putative luteal phases of each individual. Whichever feature of the four first crossed this threshold was used as the “confirmation day” of pregnancy onset (**Figure 3**).

#### Statistical analyses

To avoid assumptions of normality, feature values are reported as medians ± M.A.D. unless otherwise stated. For statistical comparisons of temperature features, Kruskal Wallis (KW) tests were used instead of ANOVAS to assess the trends in the 2 putative luteal phases, estimated based on elevation of temperature metrics, preceding pregnancy as compared to the days following conception. For KW tests, χ2 and p values are listed in the text. Dunn’s test was used to correct for multiple comparisons among luteal phase values and values of each of the first 10 days following self-reported conception. Figures were formatted in Microsoft PowerPoint 2019 (Microsoft Inc., Redmond, WA) and Adobe Photoshop CS8 (Adobe Inc, San Jose, CA).

## Data Availability

Data and code sufficient to recapitulate these results will be made available at Azure Grant's github (see in: text link)

## Acknowledgements

The authors would like to thank the individuals who approached us with pregnancy data stories over the past many years, including the QCycle study group and the Oura community. Special thanks to those at Oura who supported logistics of this study, to Dr. Eleni Greenwood Jaswa for her feedback on this manuscript, and to Lance Kriegsfeld for ongoing intellectual support of this work.

